# Predicting increases in COVID-19 incidence to identify locations for targeted testing in West Virginia: A machine learning enhanced approach

**DOI:** 10.1101/2021.10.06.21264569

**Authors:** Bradley S. Price, Maryam Khodaverdi, Adam Halasz, Brian Hendricks, Wesley Kimble, Gordon S. Smith, Sally L. Hodder

**Affiliations:** Management Information Systems Department, West Virginia University, Morgantown, West Virginia; West Virginia Clinical and Translational Science Institute, Morgantown, West Virginia; School of Mathematics and Data Science, West Virginia University, Morgantown, West Virginia; Department of Epidemiology and Biostatistics, West Virginia University, Morgantown, West Virginia; West Virginia University School of Medicine, Morgantown, West Virginia

## Abstract

During the COVID-19 pandemic, West Virginia developed an aggressive SARS-CoV-2 testing strategy which included utilizing pop-up mobile testing in locations anticipated to have near-term increases in SARS-CXoV-2 infections. In this study, we describe and compare two methods for predicting near-term SARS-CoV-2 incidence in West Virginia counties. The first method, R_t_ Only, is solely based on producing forecasts for each county using the daily instantaneous reproductive numbers, R_t._ The second method, ML+ R_t_, is a machine learning approach that uses a Long Short-Term Memory network to predict the near-term number of cases for each county using epidemiological statistics such as Rt, county population information, and time series trends including information on major holidays, as well as leveraging statewide COVID-19 trends across counties and county population size. Both approaches used daily county-level SARS-CoV-2 incidence data provided by the West Virginia Department Health and Human Resources beginning April 2020. The methods are compared on the accuracy of near-term SARS-CoV-2 increases predictions by county over 17 weeks from January 1, 2021-April 30, 2021. Both methods performed well (correlation between forecasted number of cases and the actual number of cases week over week is 0.872 for the ML+R_t_ method and 0.867 for the R_t_ Only method) but differ in performance at various time points. Over the 17-week assessment period, the ML+R_t_ method outperforms the R_t_ Only method in identifying larger spikes. We also find that both methods perform adequately in both rural and non-rural predictions. Finally, we provide a detailed discussion on practical issues regarding implementing forecasting models for public health action based on R_t_, and the potential for further development of machine learning methods that are enhanced by R_t._

## Introduction

Rural communities in the United States (US) have been heavily impacted by the novel coronavirus (SARS-CoV-2) pandemic. SARS-CoV-2 related deaths have occurred disproportionately among rural areas of the US, and negative impacts on health and economic well-being have been described to be more severe among rural populations (Bradford, Coe, Enomoto, & White, 2020) (Mueller, McConnell, & Burow, 2021) (Cyr, Etchin, & Guthrie, 2019). Persons living in rural communities often have multiple barriers to health care and laboratory diagnostic testing due to geographic, transportation, and cost.

Early in the COVID-19 pandemic, the state of West Virginia (WV) provided county-specific data on SARS-CoV-2 testing results so that daily instantaneous reproductive numbers (R_t_) could be calculated for each WV county to indicate viral transmission dynamics. An aggressive SARS-CoV-2 testing strategy was implemented that included static as well as mobile testing units. The Rapid Acceleration of Diagnostics in Underserved Populations (RADx-UP), funded by the National Institutes of Health, provided the opportunity to deliver pop-up mobile testing in those areas predicted to have the greatest increases in SARS-CoV-2 incidence. The objective was to increase testing in those communities most likely to have a near-term (within 7-10 days) increase in COVID-19 cases, thereby potentially providing early identification of SARS-CoV-2 infected persons who may then quarantine more rapidly in an effort to blunt the anticipated increase in new cases.

Two strategies to predict near-term increases in SARS-CoV-2 cases were developed using recent county-specific incidence of infections and R_t_ – one method is a dynamical algorithm-based prediction using R_t_ and the serial interval while the second method uses a long short-term memory (LSTM) machine learning strategy. The goal was to recommend counties of outbreak for targeted testing. Here we compare accuracy of the two methods to predict short-term increases in county-specific SARS-CoV-2 incidence and discuss conditions favoring one method or the other.

## Data and Methods

### Data

To obtain estimates of near-term increases in SARS-CoV-2 cases, we deployed the likelihood-based model underlying the EpiEstim package in R and developed in Cori et al. (Cori, Ferguson, Fraser, & Cauchemez, 2013) and Thompson et al. (Thompson, et al., 2019). using software provided by Imperial College London (Mishra & Valka, 2020) Two methods were employed: 1) the R_t_ Only method, a forecast based on the reproduction number and associated serial interval that predicts the future R_t_ that is then extrapolated to estimate the number of future cases; 2) a Long Short-Term Memory (LSTM) machine learning model (ML+R_t_) that utilizes the reproduction number from the R_t_ Only method as an input, but also utilizes total cases and population, among other inputs, to predict the total number of cases for a given period of time.

We received daily reports of all daily COVID-19 polymerase chain reaction (PCR) and antigen testing results conducted in WV since March 2020 directly from the WV Department of Health and Human Resources (WVDHHR). Noteworthy is that all SARS-COV-2 testing data are required to be reported to WVDHHR. Information for each unique patient is collected and contains test procurement date, test result date, patient zip code, patient county of residence, testing site name, county where the test is obtained, and test result. As patients who test positive may be tested multiple times, we only consider the first positive tests on a patient. When applying this filter, we consider data obtained from all testing sites (i.e., hospital, clinic, pharmacy, drive-through, mobile van). The number of daily cases for each county is calculated by adding the lab confirmed cases and clinical confirmed cases after filtering out repeated tests or COVID-19 diagnoses. This daily incidence data on first diagnosed infection is the basis for calculation of R_t_.

### R_t_ Only Method: Producing Short Term Predictions

Our R_t_ Only method relies on the methodology used in the EpiEstim package and the underlying modeling approach of Cori et al (Cori, Ferguson, Fraser, & Cauchemez, 2013) and Thompson et al. (Thompson, et al., 2019). This approach relates the daily incidence (number of new cases) to past cases through an instantaneous reproduction number R_t_ which characterizes the daily dynamics of transmission reflects a multitude of factors relating to individual and group behavior in the community of interest.

As a brief review, daily infections within a community occur as independent random events drawn from a Poisson distribution. The probability that exactly *k* cases occur is 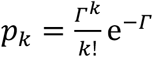, and the rate parameter Γ coincides with the average daily incidence, ⟨*k*⟩ = Γ. In the instantaneous *R*_*t*_ framework, the expected incidence on day *t* is a product of two quantities, the infection potential and the reproduction number, Γ_*t*_=Λ_*t*_*R*_*t*_. The infection potential Λ_*t*_ summarizes the record of past cases in the community and the typical variation of the infectiousness of an individual over time.

The infection potential Λ_*t*_ is determined by the incidence *I*_*t*−*s*_ on prior days *s* = 1,2, ⋯ and the serial interval distribution *w*_*s*_.

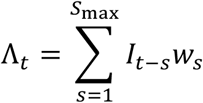

The serial interval distribution *w*_*s*_ reflects the time course of infectiousness of one infected individual at *s* = 1,2, … days from the primary infection. It encapsulates the relative increase and decrease of infectiousness of an individual, assuming all other conditions in the community remain unchanged. In practice, the serial interval is typically obtained as the normalized 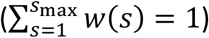 distribution of time intervals between known infector-infected pairs. Based on studies done by Gostic et al. and Challen et al., we used a distribution extending over 100 days for the serial interval (Gostic, et al., 2020) (Challen, Brooks-Pollock, Tsaneva-Atanasova, & Danon, 2020). The infection potential can be understood as the sum of the expected number of infections on day *t*, due to past cases in the community, under ideal “steady state” conditions, such that over time, each primary case causes exactly one secondary case.

The time varying reproduction number, *R*_*t*_, captures conditions of transmission that are external to the infected individuals and reflect community behavior. In this framework, *R*_*t*_ is a random variable with a Gamma distribution 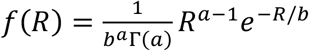. The parameters *a*_*t*_, *b*_*t*_ are determined for each day through Bayesian (maximum a posteriori probability) estimation. The parameters of interest are estimated using incidence data up to and including the current day, *I*_1_, *I*_2_, ⋯ *I*_*t*_ as follows:

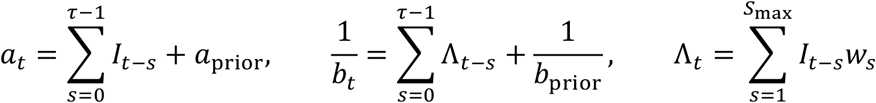

This estimated *R*_*t*_ distribution applies to the most recent *τ* days, but it requires the values of *I*_*t*′_ for *t*^′^ ≤ *t* going back to *t*^′^ = *t* − *s*_max_ where *s*_max_ is the length of the serial interval distribution. For the serial interval *w*_*s*_ we used the discretized gamma distribution with mean and standard deviation of t_s = 7.0 ± 4 days, provided in the software similar to Cori, Ferguson, Fraser, & Cauchemez (2013).

For the serial interval *w*_*s*_ we use a gamma distribution with mean and standard deviation of *τ*_*s*_ = 6.99 ± 4.02 days, as given by Flaxman, et al., 2020. Following Cori and Thompson’s method, we used a prior distribution consistent with mean and standard deviation equal to 5 (*a*_prior_ = 1, *b*_prior_ = 5).

The semi-deterministic model for future incidence, based on Cori’s method regards the daily distributions of *R*_*t*_ (values of *a*_*t*_, *b*_*t*_) as inputs that summarize the current conditions for disease transmission within the community of interest. The serial interval distribution *w*_*s*_, which is fixed with regard to time, is also an input. Thus, on day *t* we have access to the distribution of *R*_*t*_ that applies to this day (assessed using the most recent *τ* days, similar to a trailing moving average).

#### Next day prediction

Assuming we have a time series of past daily incidences {*I*_*u*_}_u=0,1,.. *t*_ ending on day *t*, the number of infections on the next day *t* + 1 follows a Poisson distribution, with parameter Γ_*t*+1_ = Λ_*t*+1_*R*_*t*+1_, where *R*_*t*+1_ is also a random variable. Assuming the parameters *a, b* of *f*(*R*_*t*+1_|*a, b*) are known, the probability of exactly *k* new infections on day *t* + 1 is:

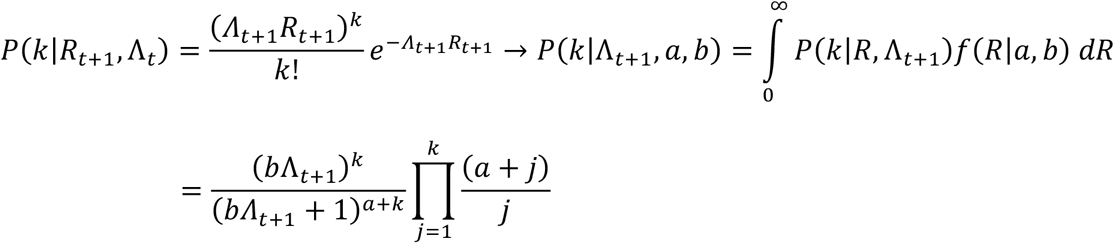

The expected number of new infections coincides with the infection potential multiplied by the expected *R*.

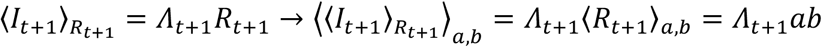

For the purpose of predicting a likelihood range for the daily incidence, we use the CDF of *R*_*t*+1_:

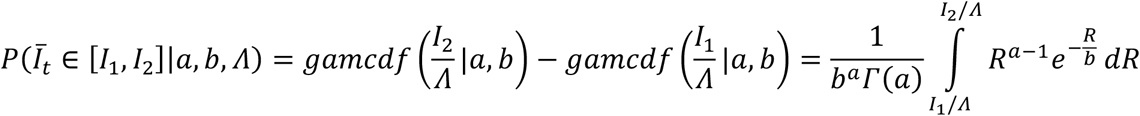

We obtain a [5% - 95%] credibility interval for the daily incidence using the inverse CDF for *R* and multiplying by the corresponding infection potential. This provides a smaller variance than the discrete distribution *P*(*k*) but is a more practical indication of the incidence rate.

#### Extrapolation over multiple days

To go beyond the “next” day, we iterate the one-day prediction, using predicted values to expand the incidence data. One can reasonably extrapolate the current distribution of *R*_*t*_ to *t* + 1 and any number of days in the future. For the short term (7 day) predictions discussed here, we assumed the value of the most recent available *R*_*t*_ remains the same over the prediction interval, 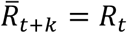.

The estimated incidence for day *t* + 1 requires the infection potential on that day Λ_*t*+1_, which is computed based on incidence up to the preceding day *t*.

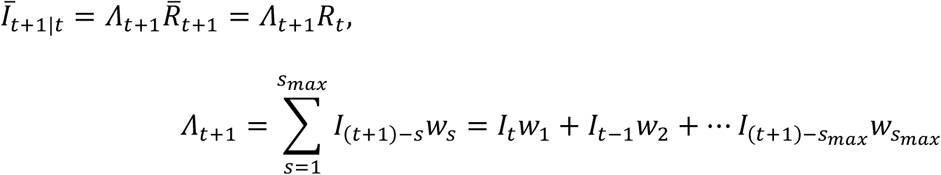

Predictions for day *t* + 2 and beyond can be obtained using the predictions for preceding days for the incidence and iteratively applying the approach for any number of k days into the future.

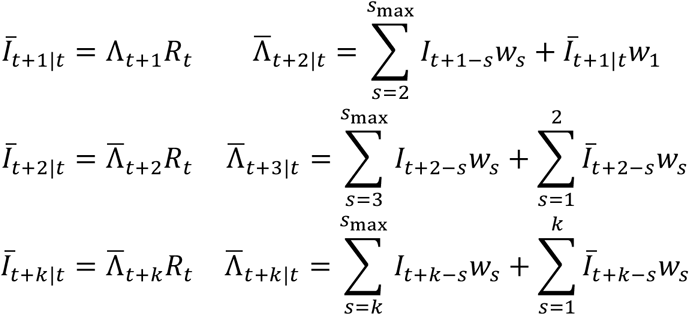

We estimate credibility intervals similar to the one-day case, using only the corresponding range for the reproduction number *R*_*t*_, and not compounding with uncertainty for each estimated incidence *Ī*_*t*+*k*_ or with the additional uncertainty due to the Poisson distribution of the daily (integer) incidence. While this provides a narrower range, the credible interval serves as a relative measure of the uncertainty affecting the prediction.

#### Correction for imported cases

Not accounting for imported SARS-CoV-2 cases into a county will lead to over estimation of R_t_. In practice, we are not able to directly identify imported cases, so an adjustment must be made to identify them. Assuming the daily incidence *I*_*t*_ can be separated into imported and community-spread parts:

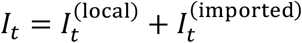

Then, imported cases are an additional input to the model. Imported cases are included in the infection potential because they contribute to new local infections, but are not included in the number of new cases when estimating the reproduction number:

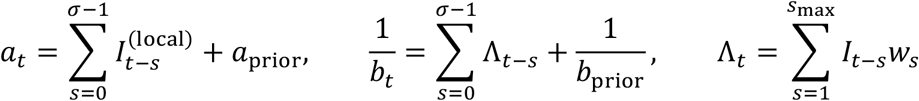

Turning to predictions, the reproduction number and infection potential computed in the standard framework can only predict the <u>local cases</u>:

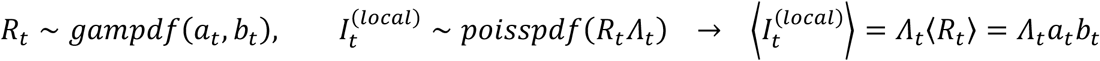

By definition, imported cases cannot be predicted in the *R*_*t*_ model; however, we can identify events when the observed number of new cases vastly exceeds the expectation from local transmission. We use this hindsight to improve our estimate of the reproduction number as follows.

We estimate the likely number of imported cases on a given day by comparing the actual incidence to the Bayesian credible interval for new local cases estimated from the previous days. This estimated past incidence is then incorporated in a corrected estimate for *R*_*t*_.

In an initial pass we compute the *a*_*t*_, *b*_*t*_ parameters for time point *t* based on the incidence time series {*I*_*τ*_}_*τ* =0,1,⋯ *t*−1)._ We compute the one-day predicted incidence on day *t* as described above, using the infection potential Λ_*t*_ and the distribution of 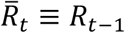 (so we do not rely on the knowledge of *I*_*t*_). We take the value corresponding to the upper *θ* = 95% credible interval as a cutoff and identify the part of the incidence that exceeds the cutoff with imported cases.

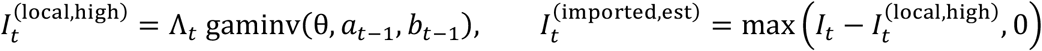

We use the estimated local incidence 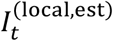 to provide revised estimates for the reproduction number as described above (also consistent with Cori and Thompson’s approach). Finally, we use the resulting *R*_*t*_ parameters for the most recent day and the full incidence to provide revised estimates for days *t* + 1, *t* + 2, … *t* + *k*.

### ML+R_t_ Method: Using Long Short-Term Memory (LSTM) Network to Forecast Outbreaks

As previously mentioned, the LSTM method implemented in this project is meant to build on the widely used R_t_ Only approach described in the previous section. The novelty of this LSTM approach is that it provides for the input of epidemiological modeling while taking advantage of cutting-edge machine learning techniques. The combination of the two allows the LSTM model to incorporate the epidemiological principles used to produce the R_t_ estimate while adding additional information such as temporal and demographic information that can be leveraged with traditional machine learning models. Further, the calculation of R_t_ using the Rt Only method uses independent data sets for each county in turn creating a unique model for each county that does not consider the impact of possible relationships between counties. By contrast, the ML+R_t_ approach uses global trends across counties. By training on all the data, we are not only able to take advantage of global trends, but by including spatial information, we are also able to show how these trends impact specific counties.

Daily county-specific R_t_, summary statistic information on the estimated R_t_ such as standard deviation, confidence intervals, and the probability of R_t_ >1 are also provided. We include values of Rt computed using both 7 and 14 day intervals. All these factors along with temporal information such as daily information on whether it is a weekend or not, holiday or not, days passed from last major holiday, days to the next major holiday were utilized as inputs for our model.

As mentioned previously, due to the length of time it takes to receive a test result (lag time), we had to consider the deflated effect on the positive cases when considering test procurement date. We observed an average lag of 3 days for results to achieve close to actual levels. To mitigate the effect of the testing lag we impute day t, t-1, t-2 with the actual SARS-CoV-2 cases for days t-3, t-4, t-5 respectively.

We utilize a Long-Short Term Memory (LSTM) recurrent neural network (Hochreiter & Schmidthuber, 1997), implemented in Python with an Adam optimizer, as our model of interest for this analysis, permitting consideration of all available county-specific input information for the past 7 days with a prediction of the number of positive cases for the county as an output. Other advantages of the LSTM approach are the ability to exploit autocorrelation between time points and the utilization of dropout layers to remove redundant information.

In general, the LSTM models are more complex versions of recursive neural networks (RNNs). The multi-layer LSTM method deployed here follows the framework described in Figure 1 where the input layer is defined by a matrix combining the number of positive cases for county c at time point t, *Y*_*c,t*_, and all inputs for county *c* at time point *t*. The inputs then move their way through the network (i.e., through the LSTM layer and dense layer) to obtain an output. The output is defined as, *Ŷ*_*c,t*+7_, the predicted daily number of cases for county *c* at time point *t+7*. LSTM can be viewed as a network where information between time points is shared. Each LSTM cell, diagramed in Figure 1, shares two pieces of information with other LSTM cells; the current state of the cell, *C*_*t*_, and output of the cell, h_*t*_, is calculated with the following formulas given input data, *x*_*t*_:

**Figure 1:** The LSTM framework deployed for the proposed ML+R_t_ method on right, and structure of each LSTM Cell on left.

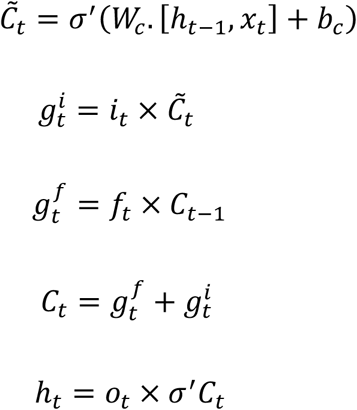

Where, *w* are the weight variables (traditionally thought of like regression coefficients), and *b* are the bias variables (traditionally thought of as intercept terms). Activation functions, *σ* and *σ*′ are non-linear transformation functions such as, sigmoid and hyperbolic tangent. A feature of each cell is input, output, and forget gates. These gates are what give the LSTM, the memory property which allows it to account and adjust for auto correlation. We define:

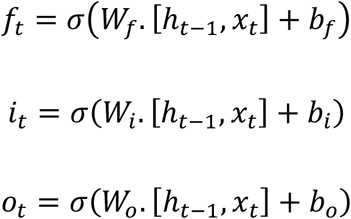

The above are gates that define the memory of the LSTM cell and are distinct linear combinations of inputs and outputs from the previous LSTM cell with specific activations functions.

In addition, as we cannot guarantee the importance of the inputs (including R_t_ and associated summary statistics), we add dropout layers which allow for the identification of important inputs. Using these dropout layers, we filtered out inputs that would be considered insignificant in order to detect the important signals coming from the input data and also protect against overfitting.

Once predictions for a given week were determined, the summary statistics of the results were produced. Summary statistics included: 1) predicted number of positive cases by county, 2) predicted percent change in cases per 100,000 persons by county compared to the previous week, 3) predicted increase in number of cases compared to the previous week, and 4) predicted number of cases relative to the population size.

### Evaluations of Models in Deployment

#### Metrics and Evaluation

To evaluate performance of the two methods, the predicted values for new SARS-CoV-2 cases were benchmarked against the actual number of positive cases recorded for each week from January 1, 2021 through April 30, 2021. As a main goal of these new case forecasts was to target areas for diagnostic testing, we viewed each week’s prediction as a recommendation. These recommendations were ranked on many several metrics but most predominately on the percentage increase in cases over the previous week. To evaluate the recommendations, we measured the total discounted cumulative gain (DCG) of each method (Järvelin & Kekäläinen, 2002). DCG is a commonly used metric in page ranking calculations and is suitable here as the information shared was used similar to page ranking calculations. As a reminder the goal of this analysis is to recommend counties of increased incidences for intervention (i.e., increased SARS-CoV-2 testing), not to predict the actual number of incidence. DCG provides a metric for comparison of differing recommendation methods, which is how both the ML+ R_t_ and Rt Only are being used. Unlike most metrics used in machine learning such as squared error or absolute error, larger DCG values indicate better performance.

To better study performance of the ML+R_t_ and R_t_ Only methods, we define two separate DCG metrics to consider the cost of poor recommendations. The first is on the ability to identify the top counties of increase regardless of the level of increase, while the second metric considers the size of the increase (percentage) in the comparison.

To define the first metric let *Ŷ*_*c,t*_ and *y*_*c,t*_ represent the number of predicted cases and actual cases over a 7-day period for the *c*th county at time point *t* respectively. To keep from biasing the evaluation towards rural areas with a low incidence, we only consider those with *y*_*c,t*+1_ > 10. Define *S*_*t*_ to be the set of indices, the largest 10 values of 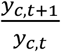 for a given time point. We defined the Binary Discounted Cumulative Gain (Binary DCG) of a set of rankings at time point *t* as:

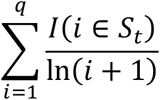

where *I*(*i* ∈ *S*_*t*_) is an indicator of a correct identification of a top 10 ranking in the actual percentage increases, and *q* is the number of rankings used in the calculation. For example, if *q* = 10, then *BDCG*_*t*_ would only evaluate the top 10 rankings, in our setting this would be the top 10 counties, returned by a method. One may view B-DCG as a weighted identifier to measure the quality of the rankings for purposes of identifying case increases (or spikes) of the top *q* recommendations.

As the closeness of the predicted number of cases to the actual case number, i.e., the “quality” of the prediction, we considered a second metric to consider the quality of the prediction rather than just considering a binary outcome. To accomplish this, we define Spike DCG as:

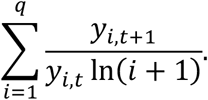

Spike DCG considers the relative size of the spike for the top *q* recommendations. While Binary DCG investigates the ability of a method to correctly identify the top 10 counties, Spike DCG places value on the recommendations that are produced by identification of larger spikes. This comparison is of great importance as targeted interventions may only have finite resources to deploy so understanding the level of trust and impact expected by the two methods is of importance.

As both the R_t_ Only and ML+R_t_ methods are used to recommend county level locations for testing, we also want to investigate the quality of the top recommendations, disregarding the order and quality of the ranked predictions. This evaluation gives a sense of the quality of the recommendations produced by the methods, relative to others.

Finally, as this study is being deployed in a state with many rural areas, we analyzed any differences in methods between rural and non-rural areas. We used the 2013 Rural-Urban Continuum Codes (RUCC) (Rural-Urban Continuum Codes (RUCC)) which define a rural area as a non-metro area with population under 20,000 and is not adjacent to an urban metro area. To asses quality of the predictions provided by each method, we examined correlations between predicted and actual 7-day positive case totals. We also assess the quality of Binary DCG and Spike DCG in both rural and non-rural areas by investigating the performance of ML+R_t_ and R_t_ Only methods among lower population communities with less access to large healthcare systems. Both R_t_ Only and ML+R_t_ methods were deployed each week from January 1, 2021 through April 30,2021 using all available training data beginning in April 2002 for each of the 55 counties in the state of WV, and resulting county recommendations were retained for comparison against the actual number of cases.

## Results

The daily number of tests from April 2020-April 2021 were highly variable (Figure 2 with some weeks having very low testing rates as illustrated by Figure 3). Each of the two prediction methods utilized all available data and was updated weekly to obtain county level predictions. We note that this study specifically focuses on evaluating predictions in the latter part of this time frame, and coincided with vaccinations becoming available to different demographics of residents of West Virginia residents, though data from the entire study was used to train each of the methods.

**Figure 2:** Number of SARS-COV-2 tests in the state of West Virginia from April 2020-April 2021.

**Figure 3:** Number of SARS-COV-2 tests in the state West Virginia from May 2020- July 2020.

The correlation between forecasted number of cases and the actual number of cases week over week is 0.872 for the ML+R_t_ method and 0.867 for the R_t_ Only method. Figure 4 shows a scatter plot of the relationship between forecasted cases and the actual corresponding cases.

**Figure 4:** A comparison of actual 7-day case totals and predicted 7-day cases totals for the ML and R_t_ methods

Figure 5 compares Binary and Spike DCG for the case of recommending 10 counties (q=10) and 55 counties (q=55). Both the R_t_ Only and ML+R_t_ methods perform well overall but differ in performance at various time points. In the case of Binary DCG the Rt Only method has better performance, and in the case of Spike DCG the ML+R_t_ method performs better. Over the 17-week assessment period, the ML+R_t_ method outperforms the R_t_ Only method in recommendations with regard to all measures except Binary DCG for q=10 (Table 1). These results show that if users are interested in mitigating outbreaks by identifying larger spikes in the Top 10 recommendations, as was the goal of this implementation, the ML+R_t_ method should be used.

**Table 1:** A comparison of total both DCG metrics for recommendations of 10 counties and 55 counties for the ML and R_t_ methods implemented.

|  |  | Binary DCG | Spike DCG |
| --- | --- | --- | --- |
| 55<br>Counties | ML+$R_t$ | 42.50 | 22.90 |
| | $R_t$ Only | 41.83 | 21.18 |
| 10<br>Counties | ML+$R_t$ | 11.88 | 7.87 |
| | $R_t$ Only | 12.59 | 4.26 |

**Figure 5:** A comparison of the ML+R_t_ and Rt Only methods with respect to Binary DCG and Spike DCG over the 17-week evaluation period for both 10 and 55 county recommendations.

A more concerning result is the decrease in both DCG metrics that are seen with regard to both methods over time. Further investigation and analysis showed that during deployment the focus of providers shifted from active testing and contact tracing to vaccination.

### Assessing Rural vs Non-Rural Results

Critically important is analysis on the performance of the two forecasting strategies in rural compared with more urban counties in WV. Correlations between predicted 7-day positive case totals and actual 7-day positive case totals are higher for non-rural counties than rural counties for both methods (Table 2).

**Table 2:** A comparison correlation of 7-day positive case totals and 7-day actual case, and both DCG metrics (total) for the ML and R_t_ methods implemented when viewed by rural and non-rural counties.

|  |  | Correlation | Binary DCG | Spike DCG |
| --- | --- | --- | --- | --- |
| Rural | ML+$R_t$ | 0.690 | 4.12 | 0.84 |
| | $R_t$ Only | 0.710 | 6.07 | 0.76 |
| Non-Rural | ML+$R_t$ | 0.867 | 7.77 | 7.03 |
| | $R_t$ Only | 0.862 | 6.52 | 3.50 |

For rural areas, the two methods perform similarly with the ML+R_t_ method slightly outperforming R_t_ Only in regard to Spike DCG (Figure 6). For non-rural areas, we observe that ML+R_t_ outperforms Rt Only for both DCG metrics (Table 2). The R_t_ Only Method performs well when identifying counties in the top 10, but ML+R_t_ method identifies larger spikes in the top 10 recommendations.

**Figure 6:** A comparison of Binary DCG and Spike DCG for both rural and non-rural counties.

A secondary analysis shows that the ML+R_t_ method recommends for enhanced SARS-CoV-2 testing more non-rural counties than rural counties in the top 10 rankings during January and February when compared to the R_t_ Only method. The opposite occurs during the March and April time period during which the R_t_ Only method recommends more non-rural counties in the top 10 compared to the ML+R_t_ methods. When coupled with decreasing number of tests, leading to lower daily incidence this alleviates any concern of bias of the ML method on rural counties.

## Discussion

In this study, we deployed two methods to predict short term incidence of SARS-CoV-2 infection for purposes of identifying West Virginia counties that might benefit from enhanced SARS-CoV-2 testing. One method, R_t_ Only, utilizes the Cori model [5], assuming that all positive cases are known. In contrast, the ML+R_t_ method utilizes R_t_ as an input value, but bases predictions on an LSTM framework that utilizes other factors such as population size.

Our results demonstrate that both methods perform well. The ML+R_t_ out performs the R_t_ only method when it comes to recommending larger spikes in the top recommendations. The implementation of the ML+R_t_ method is novel as it is utilizing epidemiological underpinnings while exploiting other information such as county population, minimum and maximum values of R_t_, variability in R_t_, and other information that may, or may not be useful in predicting out breaks.

Each of the methods for incidence prediction have strengths and weaknesses. The R_t_ Only method only assumes that all positive cases are known. However, in practice, this assumption is unreasonable and highlights some of the problems with applying the standard Cori R_t_ model to SARS-CoV-2 data. The R_t_ Only approach relies on the most recent testing data available, and our daily incidence *I*_*t*_ represents the number of positive test results from tests performed on day *t*. Publicly reported case numbers (Dong, Du, & Gardner, 2020) typically represent the number of positive test results reported on the respective day, but the lag time from test procurement varies. Using the day tests were procured eliminates one additional source of variability and brings our proxy for the “serial interval” closer to the relevant distribution (which would be the infectivity profile – see (Challen, Brooks-Pollock, Tsaneva-Atanasova, & Danon, 2020) (Britton & Scalia Tomba, 2019) (Gostic, et al., 2020)). However, this raises a practical issue in that data for day *t* is typically incomplete on day *t* and is reported gradually over several days. To address this issue, we estimate SARS-CoV-2 incidence using data from 3 days prior (*τ*_report_ = *incidence at t* − 3 days). For example, the weekly total reported on day *t* = May 12, 2021 represents the week ending on May 9, 2021, and it is this incidence that is used to predict SARS-C0V-2 incidence for the subsequent 7 days.

A second issue with the R_t_ Only method is that we do not have access to a reliable record of imported cases as they are a theoretical concept in this model. In practical settings, the term “imported” is to be taken in a (very) broad sense. There are a number of situations that have a similar effect.

1. True “exogenous” cases likely occurred due to county residents traveling for school or holidays. [1][2]. There are numerous anecdotal instances in the media but no consistent methodology or documentation of such cases. Commuters from one county to another or out of state could be susceptible to outbreaks outside of their “home” geographic area.
2. “Institutional” or “congregate setting” cases, occur (rather, are identified) over a short time in closed or limited access facilities. Congregate setting outbreaks have somewhat similar features; however, it is not obvious whether individuals infected in congregate settings (e.g., nursing homes) cause new infections in the community as these individuals have limited community access.
3. Finally, significant variability over time of test availability and policies (e.g., limited test availability early in the pandemic, prioritizing resources for vaccine rollout to the detriment of testing availability) complicates the role of the observed incidence as an estimator of the true number of infections.
4. Severity of a disease leading to hospitalization or other interventions that allows for insight into a group that was not previously being tested.

To address these issues, we use the Bayesian credible interval to better define the number of imported cases in the R_t_ Only method. By the iterative fitting technique proposed we are able to better estimate the number of imported cases that will be observed.

The ML+R_t_ value suffers from issues with practical implementation as well. The same issues with data quality from testing lags can be found when using any data driven method to forecast cases. In addition, there are known problem of using neural networks and deep learning methods when sample sizes are not extremely large. Our approach which predicts using a model that is trained from all combinations of counties and time points takes advantage of the 55 counties over the 365+ days of observed data. Early on in a pandemic it would be unreasonable to think an LSTM or many data driven methods could be used and would be reliable due to a limited number of data point. Therefore, early in the pandemic, our results show the stability of the dynamical model underlying the R_t_ Only method is reliable once the serial interval could be constructed as the Bayesian approach of the R_t_ Only method utilizes the serial interval to create an informed prior distribution of spread. For this reason, the LSTM method was not incorporated until October 2020 and only presented in this study from January through April 2021, a time period at which the SARS-CoV-2 epidemic in West Virginia was well established and just before the new Delta variant became established (only one case of Delta was identified during the study period). As the ML+R_t_ method utilizes all data available, it is less predictive during times that diagnostic testing is erratic (e.g., school breaks, testing supply shortages, etc) (Figure A1). The R_t_ Only method is able to adjust predictions in a quicker time frame Figures 3 and Figure 4 demonstrate a sharp decrease in performance of the ML+R_t_ method in February at which time there was a sharp decrease is SARS-CoV-2 diagnostic testing. Again, we recommend using the R_t_ Only approach when drastic changes in testing occur and doing so until testing stabilizes.

As we have seen during the SARS-COV-2 pandemic, situations are dynamic and models must be built to account for the changing landscape of the data and inputs available. With this in mind, extensions of this work should consider vaccination rates, population distributions, vaccine hesitancy, and baseline testing access to better predict outbreaks and target testing. A combination of vaccine information could account for decrease testing and smaller number of cases in models such as the ML+R_t_ method can adjust for this new input and do so in ways that cannot be accounted for using the Rt Only method. Furthermore, this could lead to interesting results in both identification of not only outbreaks but areas for potential variants and the possibility to use model averaging techniques to create an optimized rule that utilizes both methods.

The approaches proposed in this work provide a framework for forecasting outbreaks at a local level that utilizes two different approaches. The first is a model based on epidemiological theory, while the second is a machine learning approach that simultaneously considers historic trends and other inputs. Both methods are useful specifically the R_t_ Only method when data is limited, while the ML+R_t_ method performs well when data has been collected and a historic perspective can be presented.

## Limitations

This study addressed the West Virginia SARS-CoV-2 epidemic from January – April 2021. At that time, only one case of the Delta variant had been detected, therefore, our models do not address prediction of new SARS-CoV-2 incidence when Delta is the prevalent variant. As the Delta variant has unique epidemiologic characteristics compared to earlier SARS-CoV-2 variants such as a shortened serial interval which influences calculation of R_t_, models must be adjusted as new more virulent strains of SARS-CoV-2 appear in the population (Baisheng, et al., 2021).

## Conclusion

This study provides important information on strategies for predicting near-term increases in SARS-CoV-2 incidence, and hence, for targeting SARS-CoV-2 testing. We provide a new approach, R_t_ Only, that utilizes the estimation of the reproduction number to provide recommendations on county-specific areas where outbreaks will likely occur. We also describe a second approach, ML+R_t_, utilizing long short-term memory models that consider epidemiological statistics such as R_t_, county population information, and time series trends including information on major holidays to forecast outbreaks and create county recommendations. Comparison of the two approaches shows the top 10 recommendations produced by the ML+R_t_ method outperform the R_t_ Only method over the period of this study. Our data suggest that traditional epidemiological modeling can be enhanced by modern machine learning tools to inform decisions on where to target SARS-CoV2 testing.

## Data Availability

The data for this manuscript is protected to ensure privacy. Code will be made available upon final publication in a peer-reviewed journal.

## Acknowledgements

The project described was supported by the National Institute Of General Medical Sciences, 5U54GM104942-04 and 5U54GM104942-05S3. The content is solely the responsibility of the authors and does not necessarily represent the official views of the NIH. The authors would like to thank the West Virginia Department of Health and Human Resources, West Virginia’s Governors Joint Inter-Agency Task-Force on COVID-19 Vaccination, and Stacey Whanger. Finally, the authors would like to recognize and thank the men and women who have been on the front lines testing and treating patients during the COVID-19 pandemic.

## Appendix

### Distribution and expectation of daily incidence

The daily incidence has a Poisson distribution with parameter Λ_*t*_*R*_*t*_. *R*_*t*_ is represented as a random variable following a gamma distribution with parameters *a, b*:

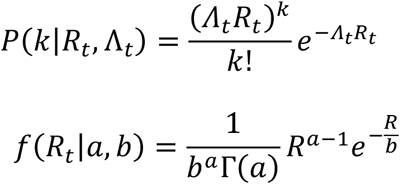

where Γ(*a*) is the usual Gamma function defined as:

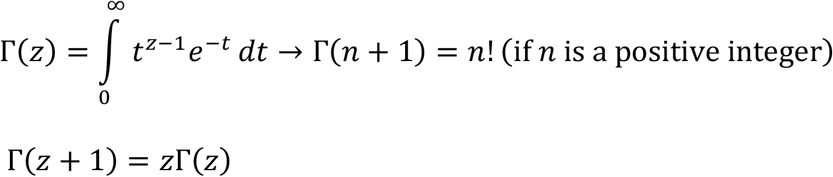

Denote by *C*_*a,b*_ the normalization constant for the Gamma distribution:

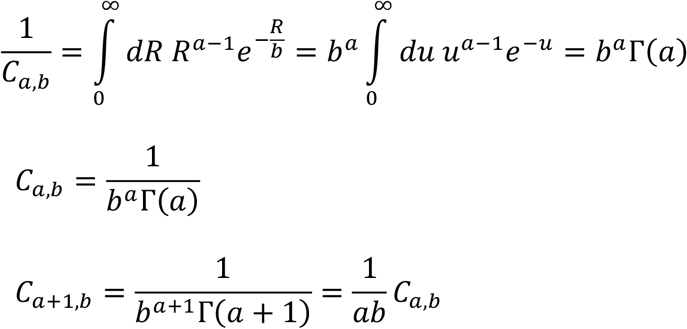

The PMF of the expected number of cases is obtained by integrating over the values of *R*_*t*_:

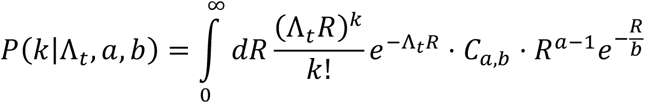

The integrand is proportional to a gamma distribution with parameters 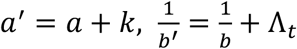

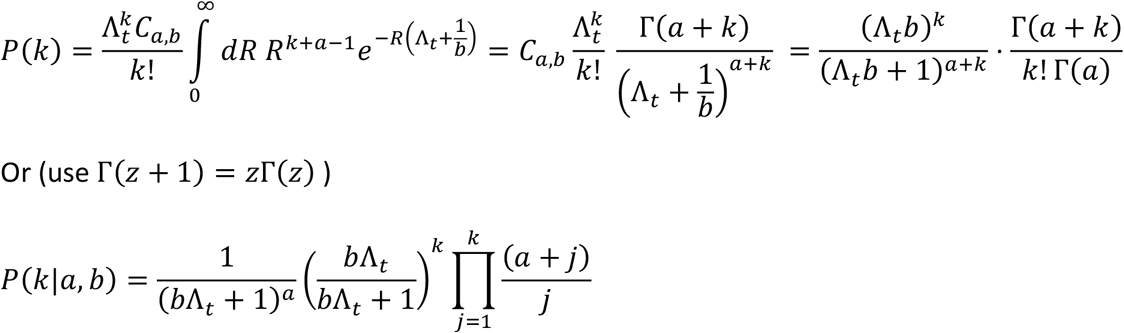

The expected number of new infections follows from working out the Gamma-Poisson distribution and coincides with the infection potential multiplied by the expected *R*

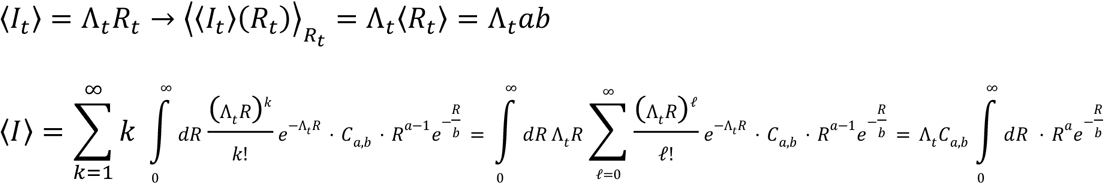

